# The burden of human coronavirus infection in children hospitalised with lower respiratory tract infection in Cape Town, South Africa

**DOI:** 10.1101/2022.07.24.22277967

**Authors:** Abdulmumuni S. Aliyu, Adelaide N. Masu, Benjamin M. Kagina, Rudzani Muloiwa

## Abstract

**Introduction:** Human coronaviruses (HCoV) NL63, HKU, OC43 and 229E are known to cause various respiratory infections including croup, pneumonia, and bronchitis in young children. The role of these four HCoV strains in the aetiology of pneumonia is little described in South Africa.

**Methods:** We used data collected between September 2012 – September 2013 from children aged <13 years with lower respiratory illness at Red Cross War Memorial Children’s Hospital. Respiratory samples including a nasopharyngeal swab (NP) and induced sputum (IS) were taken and tested for the four strains of coronaviruses using FTD33 multiplex real-time PCR.

**Results:** A total of 460 respiratory samples were analysed. Of these, 258 (56.0%) were male and 19 (4.1%) HIV infected. The median age of the children was 8 (IQR 4-18) months.

Nasopharyngeal (NP) samples were obtained from 460 children while induced sputum (IS) was not available for six children due to sample loss prior to analysis, leaving 454 available for analysis. A total of 42 (9.1%, 95% CI 6.7-12.1%) participants tested positive for HCoV in at least one of the two specimens. PCR was able to detect a total of 35 (7.7%) cases from the 454 tested IS specimens compared to 23 (5.0%) detected out of 460 NP samples.

The commonest detected HCoVs were coronavirus OC43 with 20 (4.3%) detected from either specimen followed by coronavirus NL63 or coronavirus HKU detected in 14 (3.0%) and 10 (2.2%) of positive test samples, respectively. The least common virus detected HCoV was coronavirus 229E detected in both positive test samples of one participant.

Overall HCoVs were detected in 23 (8.9%) of boys compared to 19 (9.1%) of the girls who returned a positive test; p=0.856. The overall age distribution of children with PCR detected HCoVs was similar to that of children with a negative result with median age of 10 (IQR 5-16) months and median of 8 (IQR 4-19) months, respectively; p=0.535. Prevalence of HCoV was 11/192 (5.7%), 23/153 (15.0%) and 8/115 (7.0%) in children <6 months old, 6-18 months and over 18 months respectively; p=0.008.

**Conclusion:** Children aged 6 to 18 months had double the risk of other age groups.

## INTRODUCTION

The emergence of the COVID-19 pandemic has brought into focus the role of coronaviruses as important pathogens in the aetiology of severe respiratory infection [1].Globally, respiratory tract infections are a major cause of under-five mortality [2].Specifically, a wide variety of viruses including non-severe acute respiratory syndrome (non-SARS)-related human coronaviruses (HCoVs) are responsible for 5% of all upper and lower respiratory tract infections in children below the age of five years worldwide [3]. For example, in 2010, lower respiratory tract infections caused about 5.8 million deaths in children younger than five years across the globe [4].

HCoVs 229E, NL63, HKU and OC43 are generally understood to cause mild respiratory illness in humans [6]. Evidence suggests that major outbreaks of more serious diseases are caused by SARS-CoV and Middle East respiratory syndrome (MERS) CoV [3]. SARS-CoV2, a novel strain is the newest addition to the HCoV family [5]. Meanwhile, HCoVs are considered relatively harmless, but severe infection is possible in premature infants, low birthweight babies and children with chronic underlying diseases [6]. The risk of being hospitalised with HCoV is higher among infants with chronic underlying health problems than among healthy ones [7]. However, some studies have found that HCoV-NL63 infection was often linked with more severe lower respiratory tract infection [3, 8, 9].

The four major HCoV strains are often found in association with lower respiratory tract disease involving infants and children suffering from pneumonia and bronchitis [3]. Specifically, HCoVs HKU and NL63 have been associated with more serious illnesses in the case of febrile convulsion, croup, and pneumonic illness. For this reason, children below the age of five years are often at risk of being hospitalised for respiratory tract infection caused by human coronaviruses [10]. Evidence from data collected on the individual viruses show that HCoV NL63 is more frequently associated with the development of croup compared with HCoV OC43 or HCoV 229E [11]. For example, a study by Van der Hoek et al found that at least 45% of children with HCoV NL63 infection had croup [12]. Similarly, studies by Wu et al. [13] and Han et al. [14] reported a high detection of HCoV NL63 infection in samples of children who had croup. In addition, other studies show that HCoVs such as HKU [14, 15] and NL63 [11, 16, 17, 18, 19] are frequently associated with bronchiolitis and pneumonia. A better understanding of the burden of HCoV infection as well as understanding of its contribution to viral pneumonia may inform an improvement in the management outcomes of childhood pneumonia. In addition, it has been demonstrated that more severe pneumonia is observed among infants hospitalised with respiratory disease due to detection of HCoV in respiratory samples [20]. This observation raises the possibility for inclusion of HCoV testing in routine viral panels which could have beneficial effects on describing aetiology of childhood pneumonia in future.

This study aimed to describe the burden of HCoVs infection in children hospitalised with lower respiratory tract infection at the Red Cross War Memorial Children’s Hospital (RCWMCH) over a period of one year.

## METHODS

### Ethics statement

This study is a sub-study of the parent study (HREC reference number 371/2011). However, ethical approval was sought and obtained from the FHS Human Research Ethics Committee, University of Cape Town (HREC REF: 507/2020). As the study uses data already collected by the original study, patient care was not affected. The investigator ensured that this study was conducted in full conformity with the principles set forth in the research guideline for Good Clinical Practice and the Declaration of Helsinki in its current version, whichever affords the greater protection to the participants.

### Study Design

The research follows a cross-sectional design with both descriptive and analytical elements. It utilises data that was prospectively collected over a one-year period from September 7, 2012 to September 6, 2013.

### Study Population

Infants and children below the age of 13 years who were hospitalised during the study period and presented with World Health Organization (WHO) defined acute lower respiratory tract infection as indicated by age-specific tachypnoea or lower chest indrawing [38]. Children could only be included into the study following written, informed parental consent.

Participants were excluded from the study if deemed too ill by the attending paediatrician to undergo induced sputum. Any child who had been hospitalised for a period of longer than 72 hours prior to recruitment were also excluded from the study to reduce the risk of including children who may have acquired the infection in hospital.

In order for the study to reflect the whole season, recruitment was limited to a maximum of four qualifying participants per day.

### Study Procedure

After obtaining consent, a detailed clinical examination was done noting the presence of cough, apnea, duration of symptoms and antibiotic treatment before admission. Additional history taking was conducted with the primary caregiver of the child. The child vaccination status was noted as recorded on the road to health card (RTHC). A physical examination was performed on each child and oxygen saturations measured and recorded. From each child, a nasopharyngeal aspirate and induced sputum specimen were taken for molecular diagnostic testing using PCR to test for HCoV. These data was collected by the original study but was not used for the corona sub-study.

Each child was weighed, and the child’s nutritional status was assessed using WHO weight for age Z scores [58]. The status was assessed as moderate to severely undernourished if the Z scores were lower, -2.

An ELISA test (Architect HIV Ag/Ab Combo, Abbott Diagnostics, Wiesbaden) was used to screen children with unknown status for HIV. In children younger than 18 months a positive ELISA test confirmed HIV infection (COBAS AmpliPrep/COBAS Taqman HIV-1, Roche Molecular Diagnostics, and Pleasanton, CA). For children older than 18 months a second positive ELISA using a different test method (Enzygnost Anti-HIV1/2 Plus, Siemens/Dade Behring, and Erlangen) was sufficient to confirm HIV infection. Children under 18 months of age with a positive PCR or older children testing positive on two ELISA tests were classified as being HIV infected. Infants less than 6 months of age whose mothers were HIV positive during pregnancy but whom themselves were not HIV infected were classified as HIV exposed uninfected.

The FTDResp33 multiplex real-time PCR assay was used to identify the presence of HCoV’s on NP and IS samples. In addition to other respiratory pathogens, the platform assessed for nucleocapsid protein gene on the RNA to find targets specific to coronaviruses 229E, NL63, HKU and 43.

### Statistics

Demographic characteristics were tabulated to provide a description of the study population. Percentages and 95% confidence intervals were used to depict proportions of categorical variables while medians with interquartile ranges were used to summarise continuous variables. The *χ*^2^ test or Fisher’s exact test were used to assess the strength of association between two categorical variables as appropriate. Continuous variables were compared using the Wilcoxon rank-sum test.

The description of HCoV was stratified by the type of coronavirus and the type of specimen used to detect the infection.

A significance level was set at a two-tailed P<0.05 for all analysis. Stata version 16 13 (Stata Corporation, College Station, Texas) was used to conduct all the statistical analyses.

## RESULTS

### Sociodemographic characteristic of study population

A total of 7792 children were admitted during the period in which the study was recruiting, including 987 that had respiratory illness. Of these, 460 (46.6%) participants were eligible and were enrolled into the study between September 2012 and September 2013. Of these, 258 (56.0%) were male and 19 (4.1%) HIV infected. The median age of the children was 8 months (IQR 4-18 months). The number of participants in crèche was 96 (20.9%) with 450 (97.8%) of the accompanying caregivers being mothers to their children. Prior use of antibiotics was reported in 173 (37.6%) children. Of those reporting prior use, the commonest antibiotics used alone were ceftriaxone and oral penicillin with 91 (52.6%) and 70 (40.5%), respectively. The remaining 12 (6.9%) had received a combination of penicillin and ceftriaxone or another antibiotic. The other baseline characteristics are shown in Table 1.

**Table 1:**
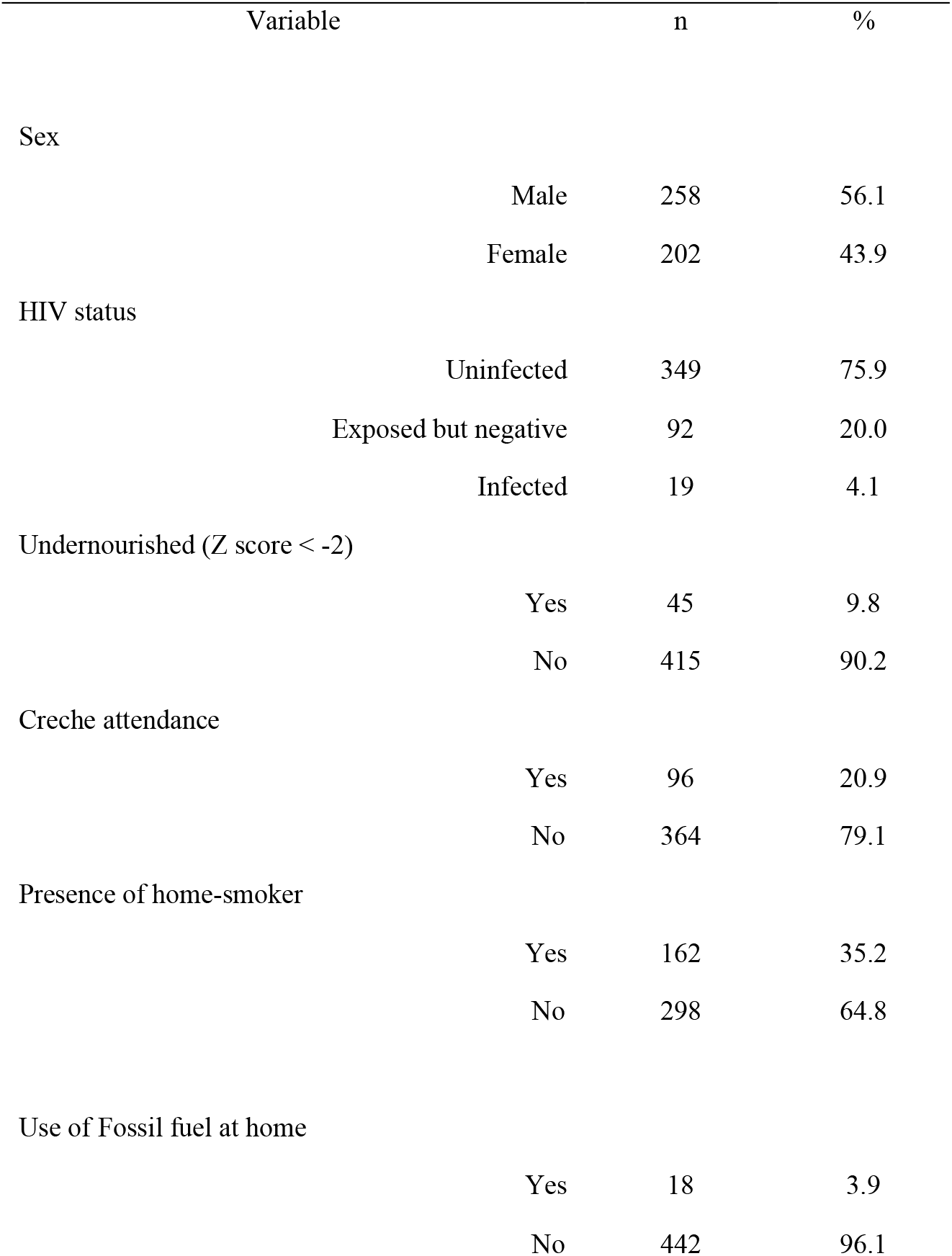
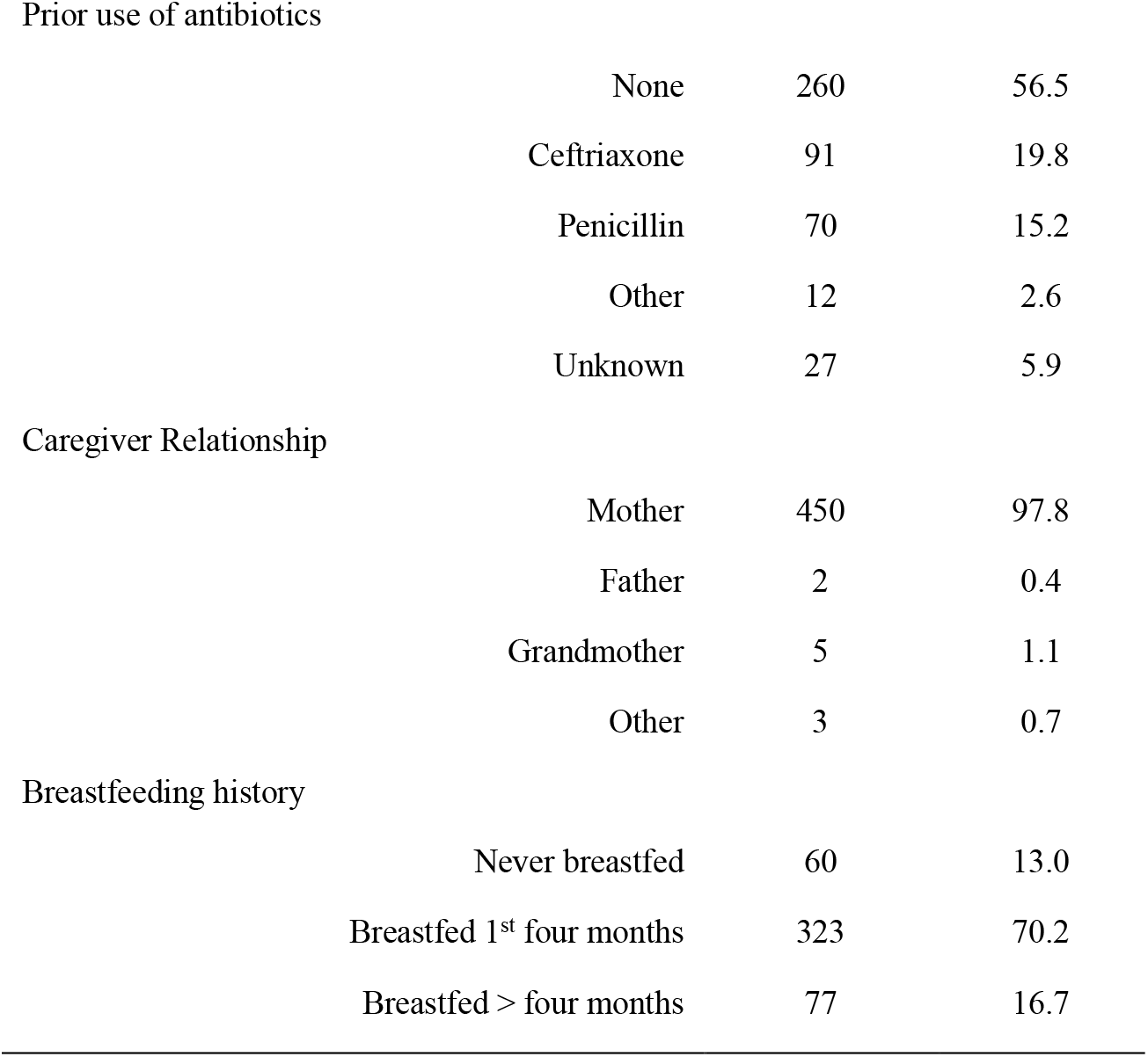
Sociodemographic characteristic of study population (N=460)

| Variable | n | % |
| --- | --- | --- |
| Sex |  |  |
| Male | 258 | 56.1 |
| Female | 202 | 43.9 |
| HIV status |  |  |
| Uninfected | 349 | 75.9 |
| Exposed but negative | 92 | 20.0 |
| Infected | 19 | 4.1 |
| Undernourished (Z score < -2) |  |  |
| Yes | 45 | 9.8 |
| No | 415 | 90.2 |
| Creche attendance |  |  |
| Yes | 96 | 20.9 |
| No | 364 | 79.1 |
| Presence of home-smoker |  |  |
| Yes | 162 | 35.2 |
| No | 298 | 64.8 |
| Use of Fossil fuel at home |  |  |
| Yes | 18 | 3.9 |
| No | 442 | 96.1 |
| Prior use of antibiotics |  |  |
| None | 260 | 56.5 |
| Ceftriaxone | 91 | 19.8 |
| Penicillin | 70 | 15.2 |
| Other | 12 | 2.6 |
| Unknown | 27 | 5.9 |
| Caregiver Relationship |  |  |
| Mother | 450 | 97.8 |
| Father | 2 | 0.4 |
| Grandmother | 5 | 1.1 |
| Other | 3 | 0.7 |
| Breastfeeding history |  |  |
| Never breastfed | 60 | 13.0 |
| Breastfed 1<sup>st</sup> four months | 323 | 70.2 |
| Breastfed > four months | 77 | 16.7 |

### Frequency of HCoV infection strains among study population by sample

Nasopharyngeal (NP) samples were obtained for all 460 children while induced sputum (IS) was not available for six children due to sample loss prior to analysis, leaving 454 available for analysis. A total of 42 (9.1%, 95% CI 6.7-12.1%) participants tested positive for HCoV in at least one of the two specimens. PCR was able to detect a total of 35 (7.7%) cases from the 454 tested IS specimens compared to 23 (5.0%) detected out of 460 NP samples. Figure

The commonest detected HCoVs was coronavirus OC43 with 20 (4.3%) detected from either specimen followed by coronavirus NL63 or coronavirus HKU detected in 14 (3.0%) and 10 (2.2%) of the participants, respectively. The least common HCoV detected was coronavirus 229E detected in both samples of one individual. Both coronaviruses OC43 and NL63 were more frequently detected on IS than NP while the other two had the same frequency in both samples (Figure 2). For the one participant, coronavirus 229E was detected in both NP and IS while coronavirus OC43 was found in both IS and NP in seven participants. Coronavirus NL63 was detected in both IS and NP in four participants, while coronavirus HKU was detected in both NP and IS in three individuals. One participant had more than one type of coronavirus in NP (coronavirus OC43 and HKU) and another one had coronavirus HKU in IS and coronavirus OC43 in NP. There were 16 participants who returned positive results and had different coronaviruses detected in both IS and NP.

**Figure 1:**
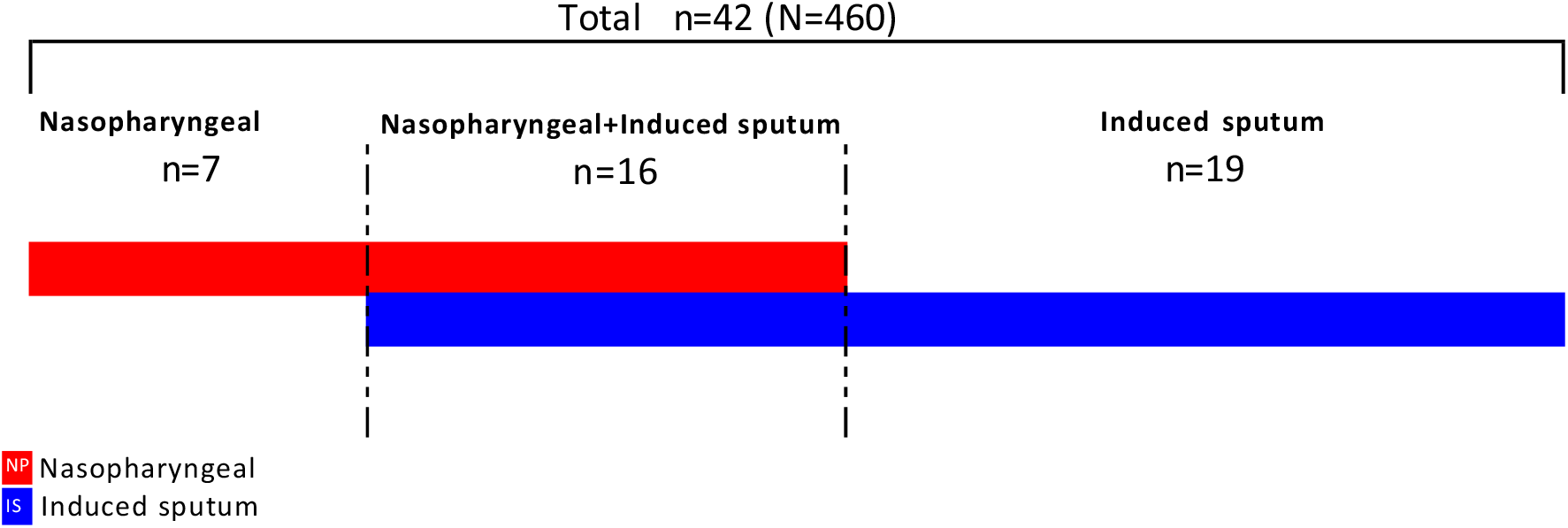
Number of positive specimens by type of sample analysed.

**Figure 2.**
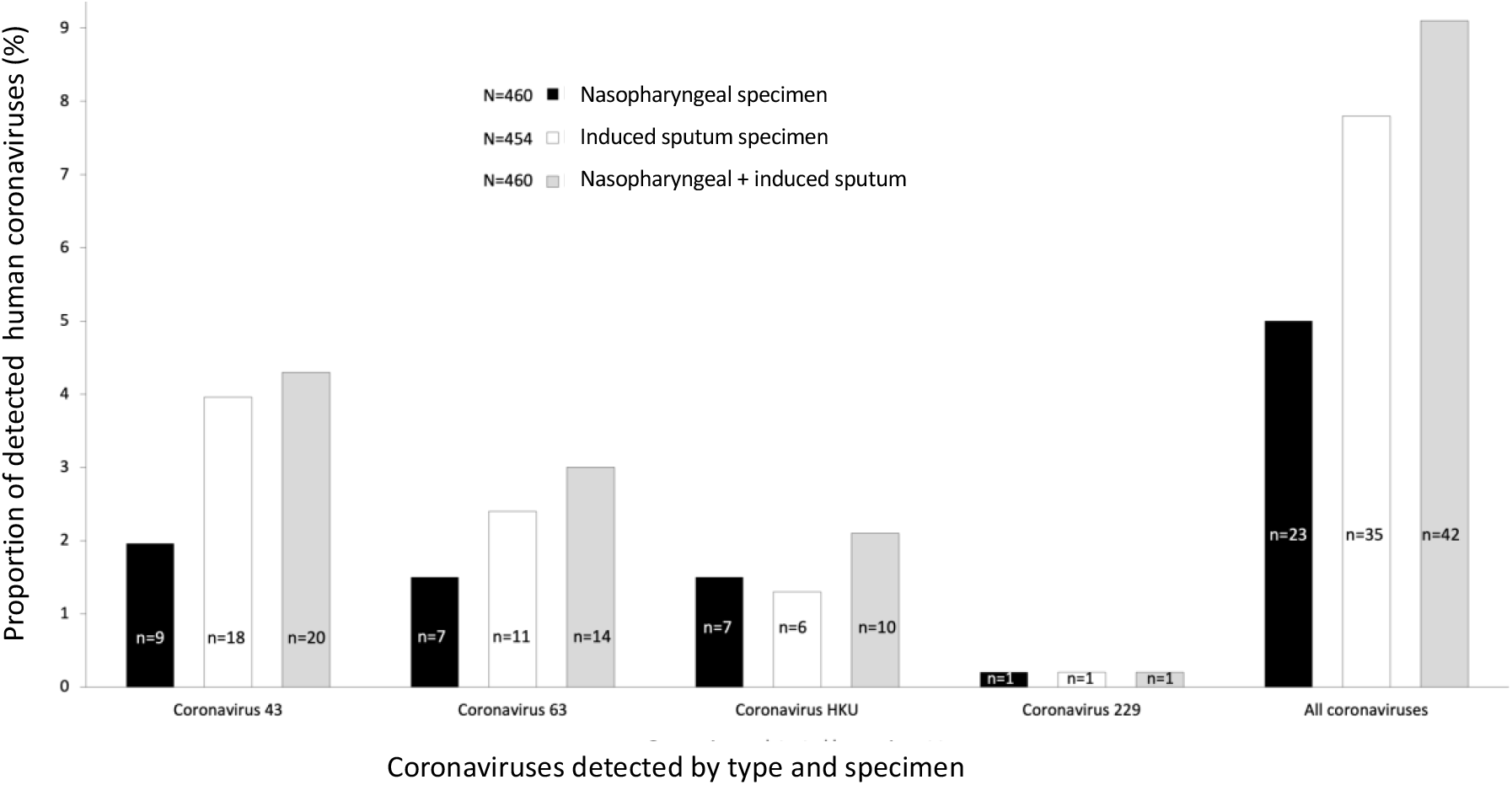
Proportion and number of detected human coronaviruses by type and sample used.

### Risk factors for HCoV infection

Overall HCoVs were detected in 23 (8.9%) of boys compared to 19 (9.1%) of the girls who returned a positive test; p=0.856. Similarly, HCoVs were detected in 30 (8.6%) of the 349 HIV unexposed uninfected children and in 12 (13.0%) HIV exposed uninfected children with none detected in HIV infected children; p=0.155. The overall age distribution of children with PCR detected HCoVs was similar to that of children with a negative result with median age of 10 (IQR 5-16) months and median of 8 (IQR 4-19) months, respectively; p=0.535. However, when the risk was stratified by age category, the frequency of children testing positive was 11/ 192 (5.7%), 23/153 (15.0%) and 8/115 (7.0%) in children less than six months of age, six to 18 months, and children above 18 months, respectively; p=0.008. Analysis of other potential risk factors is shown in Table 2.

**Table 2:**
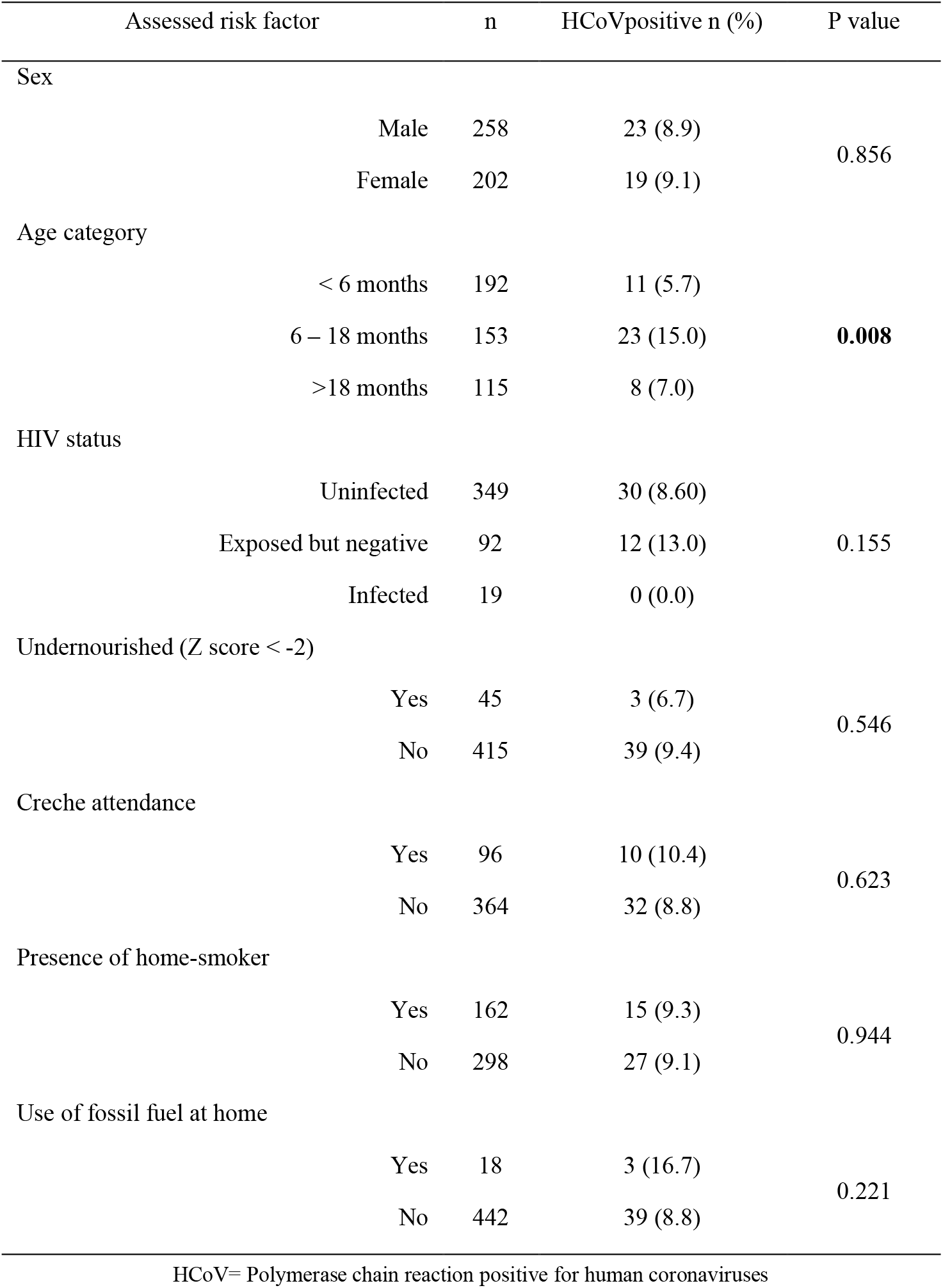
Potential risk factors for human coronavirus infection (N=460)

### Clinical presentation and outcome of HCoV infection

The commonest presentation of the study participants was cough in 456 (99.1%) with all children testing positive for HCoVs presenting with cough. Fever was found in 21 (50.0%) of HCoV positive children compared to 151 (36.1%) in PCR negative children; p=0.076. Other clinical presentations are shown in Table 3.

**Table 3.**
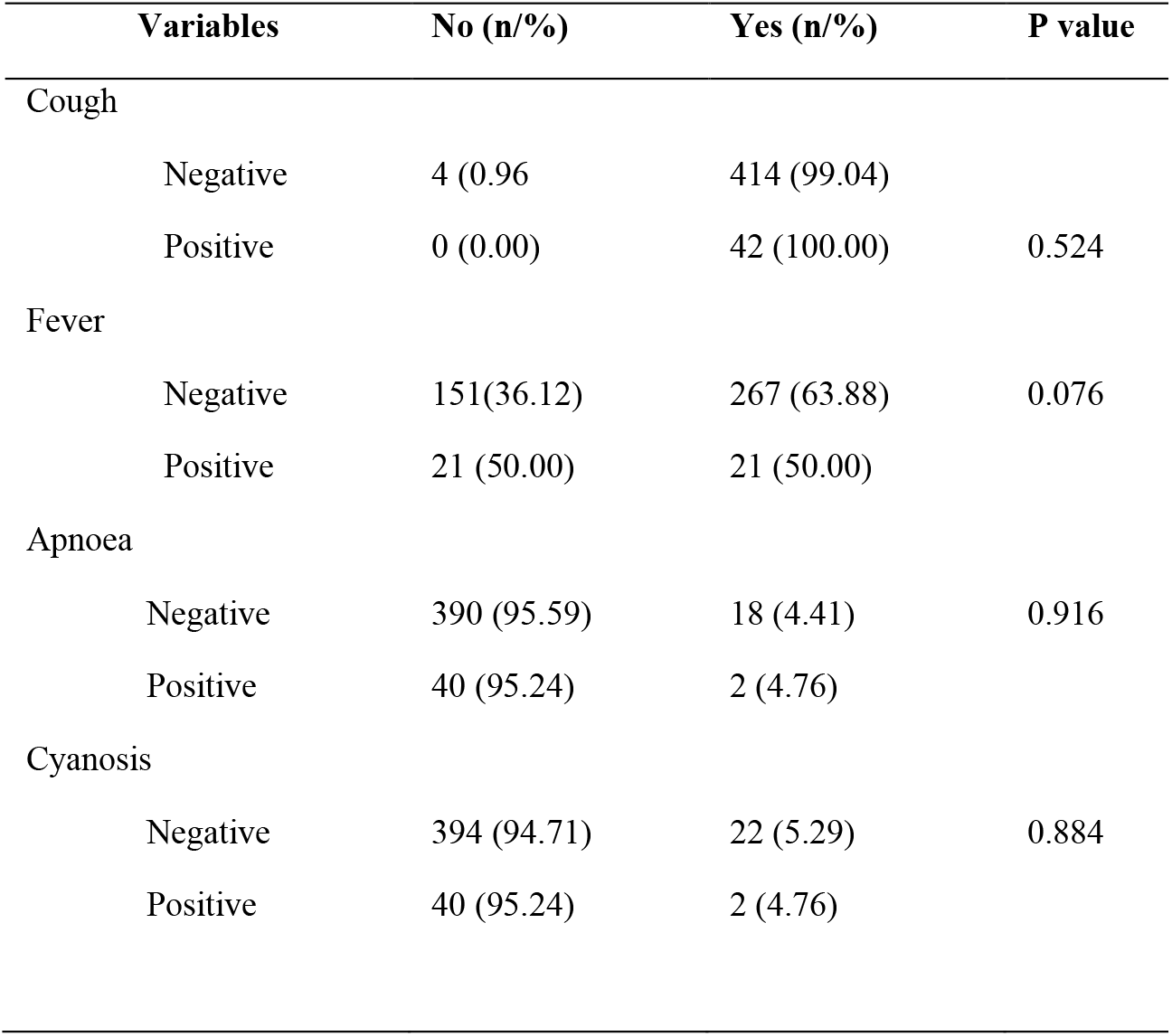
Clinical presentation of participants by coronavirus status.

There were no deaths reported in the study. The median length of hospital stay was 2 days (IQR 1 – 4) days in the group testing negative for coronaviruses compared to a median of 1.5 days (IQR 1-3) days in the group testing positive, p=0.557. Only 14 children required a high dependency care or critical care admission with one (2.4%) and 13 (3.1%) in the coronavirus positive and negative groups, respectively; p=0.629.

The median oxygen saturation of children who tested positive for coronaviruses was 97% (IQR 95 – 98%) compared to 96% (IQR 95-98%) in children who did not test positive for coronavirus; p=0.492. 7 (16.7%) out of the 42 children who tested positive for HCoV required oxygen supplementation, compared to 107 (25.5%) in the group without detected HCoVs; p=0.201.

## DISCUSSION

This study shows that human coronaviruses are common in children admitted with respiratory illness. The risk of HCoV infection was common in children between the ages of six and 18 months. In general, children with HCoVs did not present differently from children who tested negative for coronaviruses. Similarly, the clinical outcome did not differ between the two groups. Of interest, the induced sputum specimen was able to detect more cases of coronaviruses than the traditionally used nasopharyngeal specimen.

Several studies have described the detection of human coronaviruses in respiratory samples with most of them using nasopharyngeal specimen collected from patients with upper respiratory tract infection [23, 24, 25]. In this study, HCoVs NL63, OC43, 229E and HKU were tested from both nasopharyngeal (NP) swab specimen and induced sputum (IS) in infants and children with lower respiratory tract infection using a multiplex real-time PCR. Although the original study was not specifically designed to detect coronaviruses, however, our data showed that coronoviruses were common in this group of children with a prevalence of almost 10% among hospitalised children <13 years of age. The study group consisted of hospitalised patients and therefore would not have picked up children infected with coronavirus who are ill and can stay at home, or who are asymptomatic.

Using IS in addition to NP doubled the detection rate by identifying cases that otherwise would have been missed by the NP specimen alone. The Drakenstein child health study in South Africa reflect that induced sputum specimens provided better viral yield for identification of potential pathogens than nasopharyngeal swabs [1]. Our study is in keeping with this findings as it has shown that more positive cases were detected on IS compared to NP. However, in the light of COVID-19 precautions, the use of IS may not be feasible for the near future unless proper ventilation and PPE is available.

The presence of fever was higher in children testing positive for HCoV in the study although this finding was not statistically significant. This is also the experience with COVID-19 where the risk of disease in children is associated with fever. In a study done in South Korea, Chang et al found that the presence of fever was a significant parameter for predicting risk of COVID-19 disease [26]. A possible explanation for this may be the interraction between cytokines and chemokines during inflammatory response to infection and injury [27].

The study identified children between the ages of 6 and 18 months as showing a significantly higher risk of coronavirus infection that was double that of other age groups. This highlights the possible effects of waning maternal antibodies which occurs in children older than 6 months [20]. Other possible reasons include social interaction, creche attendance and effect of carriers in older children and adults [28].

Compared to RSV and other respiratory viruses in which children younger than 6 months got severe infection, vaccines have been targeted for maternal vaccination to protect them [29]. This finding is consistent with the literature in which older children who got coronavirus were reported to be less likely to get severe respiratory illness [7]. But this is not the case with older children where HCoV infection is seen to occur frequently later in life than expected. This might explain the reason why the outcome seem to be slightly better in children with HCoV infection.

The risk of HCoV infection was not associated with HIV status. This contrasts with other pathogens in published studies in South Africa in which the risk of respiratory pathogens such as RSV was shown to be associated with HIV exposure among children [20, 29]. It is not fully clear why this association with coronavirus as shown in our reported results is missing. The small sample size in which the analysis was based could be the reason why it was not detected. However, HIV remains a major problem in sub-Saharan Africa (SSA) with more children having in utero exposure to HIV even if uninfected on account of successful implementation of the prevention of mother to child transmission (PMTCT) strategy [30]. A number of studies show this group of children as a growing concern for high risk for infectious diseases [20, 30].

In our study children that come from households using fossil fuel had a higher risk to have coronavirus detected in their respiratory tract, however this finding did not reach statistical significance and does not identify fossil fuel as a risk in children. This is different from what is observed in other contexts where solid fuel generally pose a risk to children with respiratory tract infection [31]. The study found similar frequencies of HCoV infection in children who live with a smoker in their homes and those without a household smoker. In addition, HCoV may be underrepresented in this study as it included only hospitalised children, not those in the community with milder coronavirus disease.

As the study is from a secondary data analysis, it is limited by specific information that we would like to have. Although the study was sufficiently powered, it had low precision and could not demonstrate statistically significant associations. The prospective design of the study may have also limited the supposed variables that were known to predict certain risk factors such as HIV and fossil fuel. Additionally, only respiratory disease caused by HCoV was included in the study; it is not possible from the data available for this study to comment on other disease associated with HCoV.

## CONCLUSION

This study investigated the burden of HCoVs in a cohort of children hospitalised with lower respiratory tract infections. Our findings highlight important new information regarding the role of human coronaviruses in the aetiology of childhood pneumonia, but also provide explanation on the risk factor for HCoV associated pneumonia in children. The study findings further reflect on the low risk of severe disease as children are disproportionately spared even in the same household with those having the disease. It is even more important in the context of COVID-19 which poses a less risk to children. However, it is important to explore in greater detail the genomic and clinical characteristics of human coronaviruses circulating in the paediatric population as this may assist in tracking potential virulent HCoV strains capable of causing more severe illness in children.

From the review of published literature, nasopharyngeal secretions from children are widely accepted as a means of diagnosing respiratory pathogens including human coronaviruses. Although other findings exist in support of induced sputum over nasopharyngeal swabs as a better method for screening respiratory pathogens, there is no agreement regarding the choice of sample type for testing in children with lower respiratory tract infection. Moreover, should we even be testing for coronaviruses routinely if they do not seem to cause any severe disease?

## RECOMMENDATIONS

While this study which is a sub-study restricted itself to the burden of human coronaviruses in children, the main respiratory pathogen under review in the parent study was *Bordetella pertusis* which only assessed similar risk factors in the same cohort of children. There is a need to assess febrile convulsions / encephalopathy / croup / Kawasaki in patients to see the true HCoV disease burden.

In addition, although nasopharyngeal specimen is widely used for the detection of human coronavirses, this study did not report on the superiority of sampling procedures in the context of pathogen detection for young children with lower respiratory tract infection. Data conveying information about this important aspect is warranted.

## Data Availability

All data produced in the present work are contained in the manuscript

## Notes

### Competing Interest Statement

The authors have declared no competing interest.

### Funding Statement

This study did not receive any funding

### Author Declarations

The Human Research Ethics Committee of the University of Cape Town gave ethical approval for this work

